# Use of the Internet and digital devices among people with severe mental ill health during the COVID-19 pandemic restrictions

**DOI:** 10.1101/2021.06.17.21259095

**Authors:** Panagiotis Spanakis, Paul Heron, Lauren Walker, Susanne Crosland, Ruth Wadman, Elizabeth Newbronner, Gordon Johnston, Simon Gilbody, Emily Peckham

## Abstract

**Background:** Restrictions due to the COVID-19 pandemic have led to everyday reliance on digitalisation of life, including access to health care services. People with severe mental ill health (SMI – e.g., bipolar or psychosis spectrum disorders) are at greater risk for digital exclusion and it is unknown to what extent they are able to adapt to online service delivery. This cross-sectional survey study explored use of the Internet and digital devices during the pandemic restrictions and its association with physical and mental health changes.

**Methods:** 367 adults with an SMI diagnosis completed a survey (online or offline) and provided information on access to Internet connection and devices, internet skills, online activities, and barriers to using the Internet. They also self-reported changes in mental and physical health.

**Results:** During the pandemic restrictions 61.6% were limited or non-users of the Internet. The majority had access to the Internet and digital devices but around half reported knowledge deficits. Most common activities were accessing information and entertainment (88.9%), staying in touch with friends and families (84.8%), and purchasing goods (other than food) (84.3%). Most common barriers were finding the Internet ‘not interesting’ (28.3%) or ‘too difficult’ (27.9%), as well as ‘security concerns’ (22.1% to 24.3%). Using the Internet ‘a lot’ (vs ‘just a bit or not at all’) during the pandemic was associated with younger age (Adj ORs = 4.76 – 6.39, *Ps* < .001), having a diagnosis of bipolar disorder (compared to psychosis; Adj OR = 3.88, *P* < .001), or reporting a decline in mental health (compared to no decline; Adj OR = 1.92, *P* = .01).

**Conclusion:** Most people with SMI were limited or non-users of the Internet during the pandemic, which seems to be mainly attributable to lack of interest and skills, rather than lack of devices or connectivity. Older adults with psychosis should be the focus of interventions to support digital engagement in people with SMI.

## INTRODUCTION

The COVID-19 pandemic led governments in many countries, including the UK, to impose restrictions in movement and social contact, to reduce the spread of the virus (Alfano and Ercolano, 2020;May, 2020). With travelling and face-to-face activities severely disrupted, people became more reliant on the Internet to perform daily activities such as keeping in touch with loved ones and accessing support (e.g., health services and purchasing essentials) (Lloyds Bank, 2020;The Office of Communications [OfCom], 2020b). In the UK’s National Health Service (NHS), the pandemic restrictions led to a shift from traditional face to face care to remote (telephone or video call) care, both in mental health (Chen, Jones, Underwood, Moore, Bullmore, Banerjee, et al., 2020; Johns, Tan, Burhouse, Ogonovsky, Rees and Ahuja, 2020;Johnson, Dalton-Locke, Vera San Juan, Foye, Oram, Papamichail, et al., 2021) and the broader sector (Mehta, Yates, Smith, Henderson, Winteringham and Burns, 2020;Nune, Iyengar, Ahmed and Sapkota, 2020;Shah, Thakrar, Visvanathan and Thamban, 2021).

However, sizeable sections of the UK society are either non-users (13%, ca. 8.7 million people, Ofcom2020a) or only limited users of the Internet (using the Internet infrequently and for a small range of activities; 14.3%, ca. 7.4 million people, Good Things Foundation and Yates, 2017), with the main barriers being lack of access to the Internet and digital devices, lack of skills, or lack of motivation (Lloyds Bank, 2020). During the pandemic, this might lead to digital exclusion via restricted or no access to online services and activities. For example, during the early phases of the pandemic, people who were considered at risk for severe complications from COVID-19 were sent a letter containing multiple web links to sources of support. However, it was estimated that around 150,000 to 175,000 people sent this letter did not have access to the Internet (Mathers, Richardson, Vincent, Joseph and Stone, 2020).

Worryingly, vulnerable groups such as older adults, and disabled or displaced people, are more likely to be digitally excluded (All Party Parliamentary Group on Social Integration [APPG], 2020;Office for National Statistics [ONS], 2019). One such group that has traditionally faced profound inequalities are people with severe mental ill health (SMI) such as psychosis spectrum and bipolar disorders; despite this they have received very little attention in terms of digital exclusion risk. People with SMI are likely to need to attend regular health care appointments to monitor their health conditions. Often, they suffer from long-term physical illnesses leading to reduced life expectancy (Dickerson, Origoni, Rowe, Katsafanas, Newman, Ziemann, et al., 2021;Hayes, Marston, Walters, King and Osborn, 2017) and, therefore, they have been more likely to self-isolate for long periods of time during the pandemic. The pandemic restrictions were also likely to exacerbate feelings of loneliness that were already common among people with SMI prior to the pandemic (Badcock, Adery and Park, 2020). In this context, use of the Internet might have been vital for people with SMI to access health care and support for their physical and mental health needs, as well as informal social support and information during the pandemic restrictions.

Pre-COVID data shows a mixed picture of digital engagement in people with SMI. For example, a study of people with bipolar disorder found a high prevalence of digital device ownership (92.8% owning a smartphone; Hidalgo-Mazzei, Nikolova, Kitchen and Young, 2019). In a longitudinal study of people with psychosis, digital exclusion had reduced from 30% to 18.3% over 5 years but this is still a large minority (Robotham, Satkunanathan, Doughty and Wykes, 2016), and a wide divide still existed in daily internet use (56% vs 78% in the general population). It has been noted that rates of digital exclusion were much higher in people using community rehabilitation services and thus more profoundly affected by their SMI condition (only 14.4% were Internet users; Tobitt and Percival, 2019).

Although some of these findings are encouraging, it is currently unknown whether people with SMI have adapted to the increased digitalisation of life and remain connected to their sources of support. To address this knowledge gap, the OWLS-COVID 19 survey explored the digital experiences of people with SMI during the pandemic restrictions. The aim was to identify the extent to which people with SMI have been using the Internet, whether socio-demographic and health characteristics have any influence on this, and whether Internet use was associated with changes in mental or physical health. We also sought to understand what people have been using the Internet for and what barriers exist to this.

## METHODS

### Design and procedure

The Closing the Gap (CtG) study is a large clinical cohort (N = 9,914) comprising adults (aged 18 years or older) with documented diagnosis of schizophrenia or delusional/psychotic illness (ICD 10 F20.X & F22.X or DSM equivalent) or bipolar disorder (ICD F31.X or DSM equivalent) recruited between April 2016 and May 2020 (Mishu, Peckham, Heron, Tew, Stubbs and Gilbody, 2019). The Optimising Wellbeing in Self-Isolation study (OWLS) recruited a sub-cohort from CTG from July 2020 to December 2020, to explore the effects of the COVID-19 pandemic restrictions on people with severe mental ill health (owlsresearch.york.ac.uk). A purposive sampling method was used based on gender, age, ethnicity, and care setting (primary or secondary care), trying to recruit as many participants as possible from each category.

To be eligible for invitation to OWLS, CtG participants had to have provided contact details and consented to be contacted again, as well as been originally recruited from a clinical site with capacity to take part in OWLS. Out of all the eligible participants, a purposive sub-sample was selected and invited to OWLS by phone or letter. The invited sub-sample was selected based on locality within the participating sites and time of recruitment to CtG. Locality was used to provide geographical diversity, inviting participants from 17 mental health trusts and six CRN areas in England, including a mix of rural and urban settings. To increase the chances of participants responding and their contact details remaining current, we preferentially invited the participants who had been more recently recruited to the CtG cohort. Ethical approval was granted by the Health Research Authority Northwest – Liverpool Central Research Ethics Committee (REC reference 20/NW/0276).

Participants who expressed an interest in taking part in the study were provided with an information sheet (read over the phone, or send by email, text message, or post). Those consenting to participate were given the option to complete the survey off-line (over the phone or on a hard copy survey sent by post) or online, to minimise digital confidence bias in the sample.

### Measures

#### Sample characteristics

In the CtG study, participants provided information on their date of birth and ethnicity. Date of birth was used to calculate participants’ age at the time they took part in OWLS and ethnicity was coded as White or Other than White. Participants’ health records were inspected to obtain their SMI diagnosis, which was then categorised into psychosis (including schizophrenia, schizoaffective or any other psychotic disorder), bipolar disorder, or other SMI. For those not providing consent to access their records or insufficient identifiable information (e.g., name and date of birth), diagnosis was coded as “not recorded”.

In the OWLS study, participants who reported currently receiving support from mental health services were coded as secondary care patients, while those who were not receiving support from mental health services were coded as primary care patients. Participants also reported their financial situation since the beginning of the pandemic as “better”, “worse”, “about the same” or “don’t know”. After excluding those not knowing, a binary variable was derived (decline or no decline in financial situation).

#### Use of the Internet and digital technologies

Self-reported knowledge of the Internet and barriers to using the Internet were assessed using items from the Oxford Internet Survey 2019 (available at https://oxis.oii.ox.ac.uk/). Self-reported knowledge was rated by participants from 0 (poor) to 5 (outstanding) and categorised as bad, poor/fair, or good/outstanding.

Participants who reported not using the Internet or using it ‘just a little’ during the pandemic, were asked to indicate a reason for this, choosing as many as applied from a pre-specified list of barriers (e.g., I am just not interested, or it is too difficult to use). On advice of the OWLS Lived Experience Panel three of the original items from the Oxford Internet Survey were removed or adapted. For example, the item reading “it is not for people like me” was removed (language could have been perceived as stigmatising), and two items about data security concerns (I worry about being conned or having money stolen; I worry about having my personal details stolen) were rephrased into a single item using milder language (I worry about the security of my data and information) to reduce any potential triggers for psychotic symptoms (e.g., paranoia).

Participants also answered a series of single-item bespoke questions, created for the OWLS survey. These explored the following topics:

a. Access to digital devices (Do you own any of the following devices?), where participants could choose among smartphones, tablets, or laptop/desktop computers, and access to the Internet (Can you access the internet from your home – yes/no).
b. Interest in learning about the Internet (Would you like to learn more about how to use the Internet to do some your daily activities?), with response options being: There might be things I don’t know and I would be interested in learning; there might be things I don’t know but I am not interested in learning; and I already know how to do the things I want.
c. Use of the Internet during the pandemic (In general, have you used the Internet during the pandemic restrictions to do some of your daily activities - e.g., buy groceries, pay bills, etc.?), with response options being: Yes a lot, Yes a little, or No. This was then categorised into users of the Internet (yes a lot) and limited / non-users of the Internet (yes a little, no).

Participants who reported using the Internet (either a lot or a little) during the pandemic restrictions were asked to indicate the specific activities they performed, choosing as many as applied from a pre-specified list of activities.

#### Self-reported changes in physical and mental health

Participants were asked how their subjective health has changed compared to before the pandemic restrictions, with the response options being: about the same; worse than before; better than before; I do not know. This was asked for physical and mental health separately. After removing those responding that they did not know, two separate binary variables were derived coded as decline in physical/mental health (including those reporting worse that before) or no decline (including all other options).

#### Wellbeing

Wellbeing was measured with the four items used by the Office of National Statistics (Ofcom 2018), assessing life satisfaction, sense of worthwhileness, and feelings of happiness and anxiety. Each item was rated on a scale from 0 (not at all) to 10 (completely) and a total score (0-40) was calculated after reversing the last item. Higher scores indicated greater sense of wellbeing.

### Analysis

The analysis plan was pre-registered in OSF (available at https://doi.org/10.17605/OSF.IO/E3KDM - section 2.3). A complete cases analysis was performed. We used two binary logistic regression models where Internet use was the outcome variable. The first examined which sample characteristics were associated with Internet use and included age, ethnicity, socio-economic deprivation, treatment setting and diagnosis. The second examined whether Internet use was associated with wellbeing, and changes in mental and physical health. Associations of all independent variables and Internet use were examined with univariate models before added into the multivariate models. All independent variables were inserted into the multivariate model at once.

For the diagnosis variable, we deviated from our pre-registered plan by adding the “not recorded” category. This was to retain in the analysis the 48 participants that did not provide consent or sufficient details to retrieve their diagnosis from their health records.

This paper also presents a post-hoc exploratory analysis to investigate one of the findings derived from the pre-registered analysis. The post-hoc analysis examined the association of changes in mental health with use of the Internet, after adjusting for age and diagnosis.

## RESULTS

### Sample

Out of 2,932 participants in the CtG study that were eligible to be invited to OWLS, we selected a purposive sub-sample of 1,166 (39.8 %) participants and successfully contacted 688 (59%). The survey was completed by 367 participants (31.5% of those eligible to be invited and 53.3% of those successfully contacted) (Figure 1).

**Figure 1:**
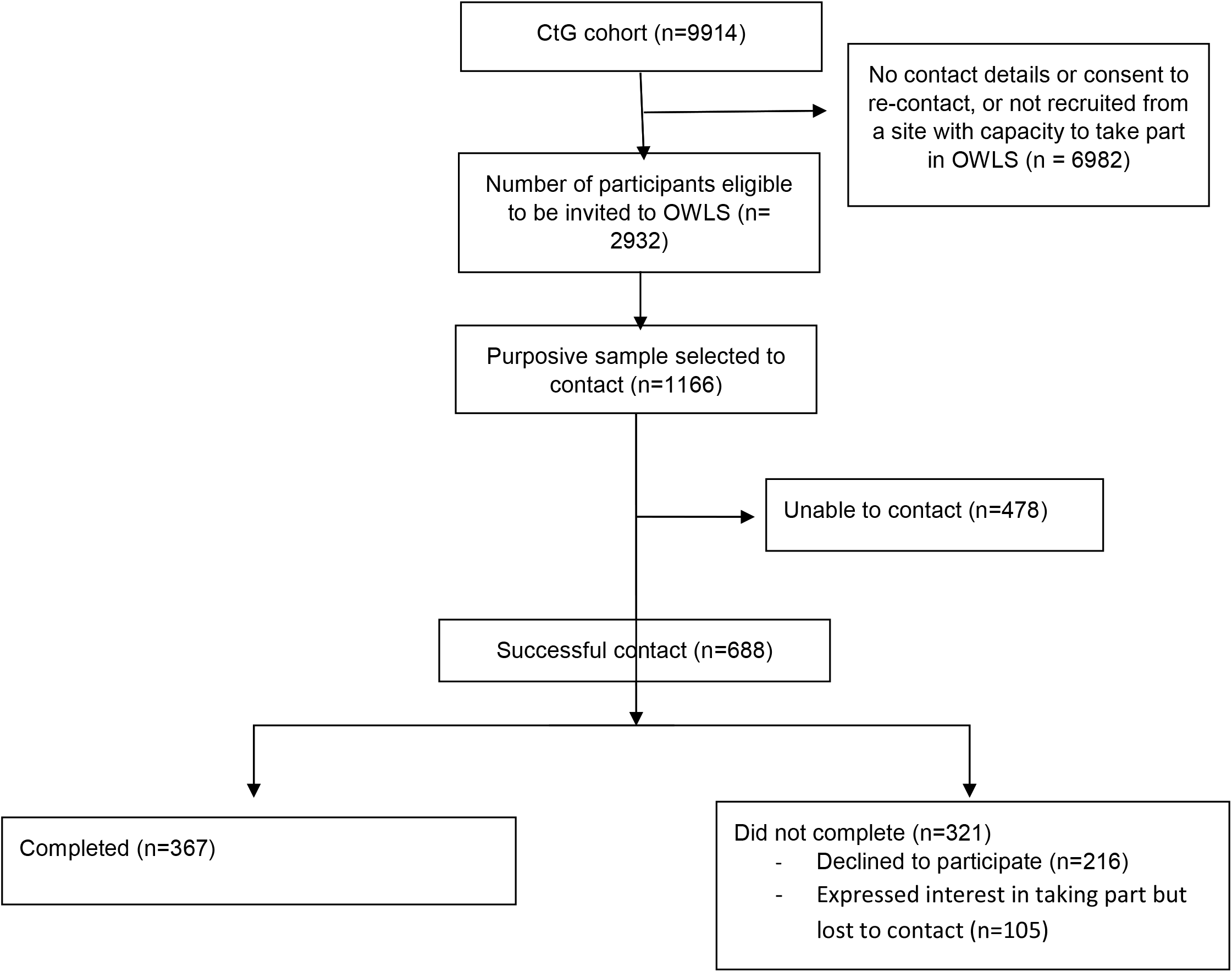
Flow Diagram - OWLS.

The final study sample (N = 367) had a mean age of 50.5 (± 15.69) years old and it included 51.0% men, 47.4% women, 1.6% transgender, 17.7% people from other than White ethnic background and 48.5% residing in high/very high deprivation areas in the country (Table 1). The primary diagnosis was psychosis (51.2%). The survey was completed online by 121 participants (33%) and over the phone or via the post by 246 (67%).

**Table 1.** Sample characteristics and health variables (N = 367)

| **Variable** | **N (%)<sup>a</sup>** |
| --- | --- |
| **Age** |  |
| 18-30 | 53 (14.4) |
| 31-45 | 97 (26.4) |
| 46-55 | 136 (37.1) |
| 66+ | 81 (22.1) |
| **Gender** |  |
| Male | 187 (51.0) |
| Female | 174 (47.4) |
| Transgender | 6 (1.6) |
| **Ethnicity** |  |
| Other than White | 65 (17.7) |
| White | 302 (82.3) |
| **Socioeconomic deprivation** |  |
| Very high | 97 (26.4) |
| High | 81 (22.1) |
| Medium | 67 (18.3) |
| Low | 55 (15.0) |
| Very low | 52 (14.2) |
| **Decline in financial situation during the pandemic** |  |
| Yes | 61 (16.6) |
| No | 285 (77.7) |
| **Mental health care setting** |  |
| Primary care | 139 (37.9) |
| Secondary care | 224 (61.0) |
| **Diagnosis** |  |
| Psychosis | 188 (51.2) |
| Bipolar disorder | 108 (29.4) |
| Other SMI | 23 (6.3) |
| Not recorded | 48 (13.1) |
| **Decline in mental health in the pandemic** |  |
| Yes | 148 (40.3) |
| No | 210 (57.2) |
| **Decline in physical health in the pandemic** |  |
| Yes | 118 (32.2) |
| No | 236 (64.3) |

a. Percentages are out of total N = 367.

### Digital engagement characteristics (Table 2)

During the pandemic restrictions, 136 participants (37.1%) were Internet-users, while 226 (61.6%) were limited or non-users of the Internet.

**Table 2.** Digital engagement characteristics (N = 367) a. Percentages are out of total N=367. b. Percentages are out of N = 182 who identified a knowledge gap.

|  | N (%)<sup>a</sup> |
| --- | --- |
| **Self-reported Internet knowledge** |  |
| Outstanding/Good | 179 (48.8) |
| Fair/Poor | 129 (35.1) |
| Bad | 39 (10.6) |
| **Device ownership** |  |
| Tablet/Smartphone | 293 (79.8) |
| Laptop/Desktop | 207 (56.4) |
| No device | 49 (13.4) |
| **Internet access at home** |  |
| Yes | 308 (83.9) |
| No | 54 (14.7) |
| **Would you like to learn more about the internet?** |  |
| **Already know what I need** | 178 (48.5) |
| **There are things I do not know** | 182 (49.6) |
| I am interested in learning more | 108 (59.3)<sup>b</sup> |
| I am not interested in learning more | 74 (40.7)<sup>b</sup> |
| **Internet use during the pandemic** |  |
| No | 145 (39.5) |
| A little | 81 (22.1) |
| No or a little (combined) | 226 (61.6) |
| A lot | 136 (37.1) |
| **Survey completion mode** |  |
| Online | 121 (33.0) |
| Phone/By post | 246 (67.0) |

Most of the participants owned a digital device and had access to the internet from home. Around half rated their knowledge of the Internet as good or outstanding and reported no knowledge gap (‘*I already know what I need*’). Of those reporting a knowledge gap, 59.3% were interested in learning more about the internet (Table 2).

The most common activities that participants used the Internet for during the pandemic restrictions were to access information or entertainment (88.9%), stay in touch with friends and family (84.8%) and purchase products other than food or groceries (84.3%). The least common activity was to stay in touch with colleagues from work (33.6%) (Figure 2).

**Figure 2.**
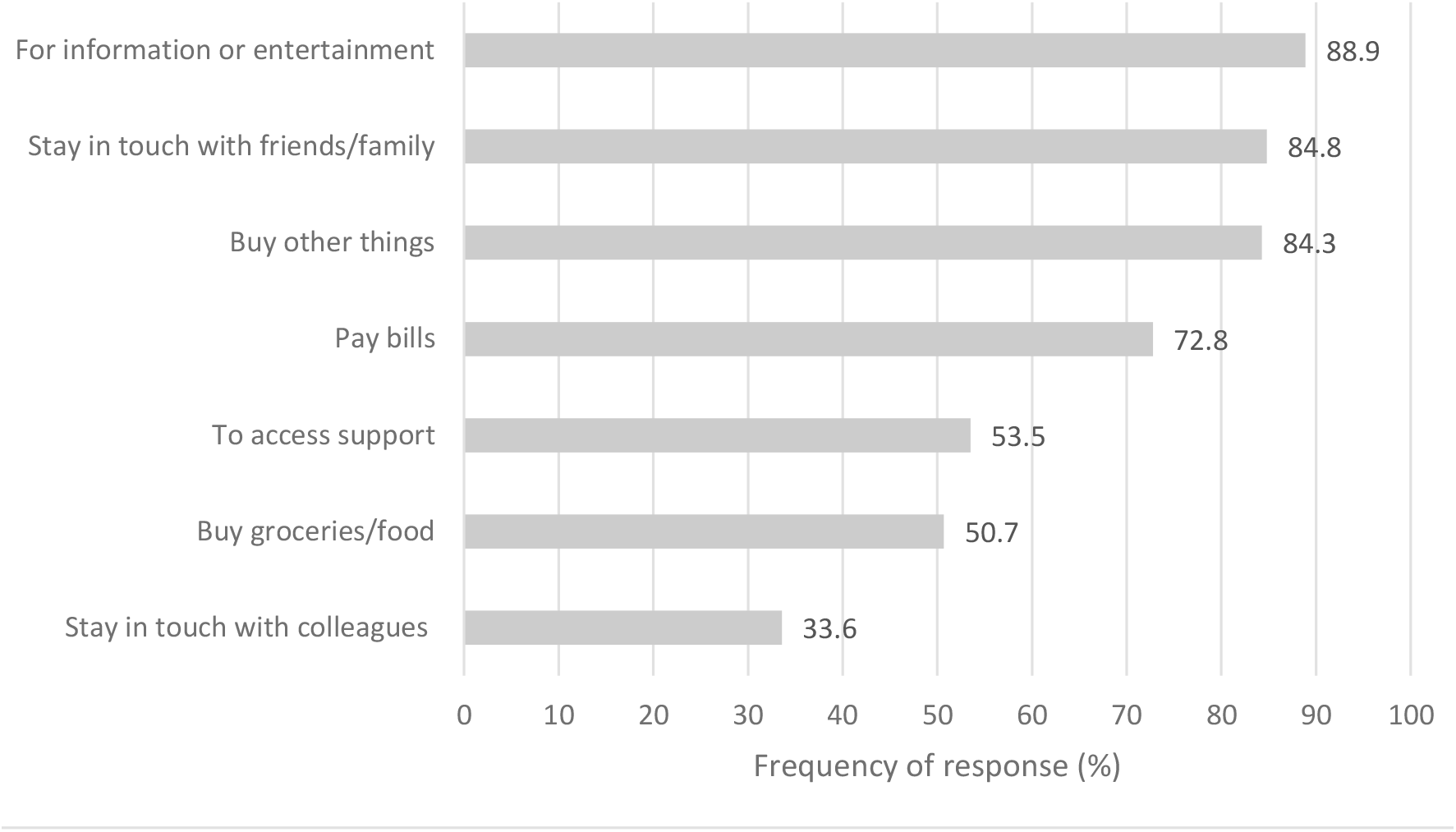
Activities performed online during the pandemic restrictions, among Internet users (limited or regular) (N = 217)

Among limited or non-users of the Internet, the most common barriers were lack of interest in using the Internet (28.3%), finding the Internet too difficult to use (27.9%), being concerned about the security of their data and information (24.3%) and being worried about their privacy (22.1%). The least reported barrier was finding the Internet not useful (3.1%) (Figure 3).

**Figure 3.**
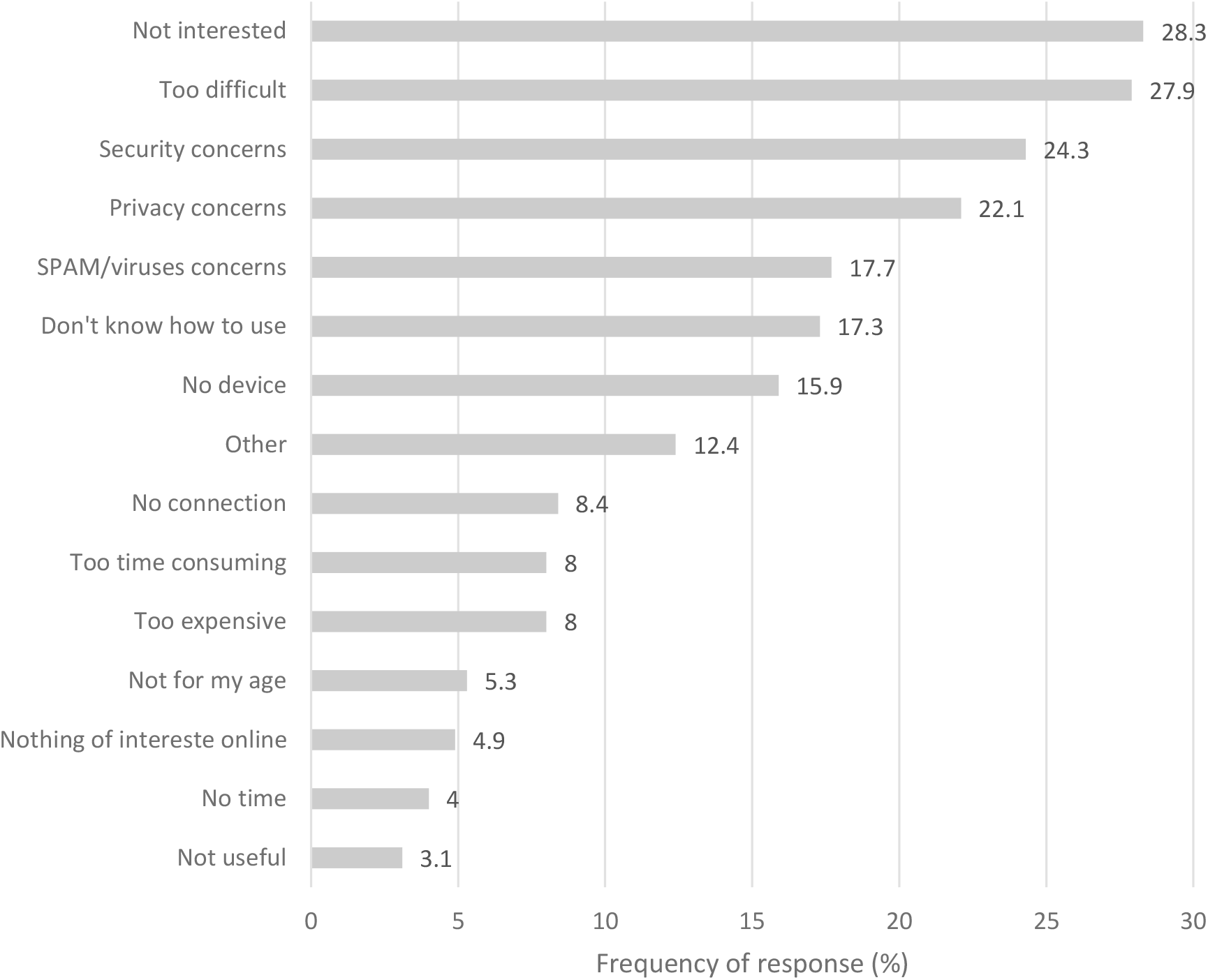
Barriers for using the Internet among limited or non-users of the Internet (N = 226)

### Associations with Internet use

In the adjusted model, younger adults (18-30 and 31-45) were five to six times more likely to have used the Internet ‘a lot’ during the pandemic, compared to older participants (66+). Gender, ethnicity, and socioeconomic deprivation were not significantly associated with use of the Internet. Participants with a diagnosis of bipolar disorder were almost four times more likely to have used the Internet a lot during the pandemic, compared to those with a psychosis-spectrum diagnosis (Table 3).

**Table 3.** Sample characteristics associated with Internet use.

|  | Using the Internet “a lot” - % (N)<sup>a</sup> | Univariate Model |  | Multivariate Model |  |
| --- | --- | --- | --- | --- | --- |
|  |  | OR | 95% CI | Adj. OR | 95% CI |
| **Age** |  |  |  |  |  |
| 18-30 | 54.9 (28) | 3.59* | 1.70 – 7.60 | 6.39** | 2.63 – 15.55 |
| 31-45 | 46.4 (45) | 2.55* | 1.34 – 4.87 | 4.76** | 2.22 – 10.20 |
| 46-65 | 31.9 (43) | 1.38 | 0.74 – 2.57 | 1.94 | 0.94 – 4.00 |
| 66+ | 25.3 (20) | 1 |  | 1 |  |
| **Minority status** |  |  |  |  |  |
| Non-BAME | 37.2 (111) | 0.93 | 0.53 – 1.61 | 0.79 | 0.41 – 1.50 |
| BAME | 39.3 (25) | 1 |  | 1 |  |
| **Deprivation** |  |  |  |  |  |
| Very high | 31.3 (30) | 0.62 | 0.31 – 1.25 | 0.61 | 0.28 – 1.35 |
| High | 38.8 (31) | 0.86 | 0.42 – 1.76 | 0.91 | 0.40 – 2.07 |
| Medium | 40.9 (27) | 0.94 | 0.45 – 1.97 | 1.15 | 0.50 – 2.64 |
| Low | 44.4 (22) | 1.09 | 0.51 – 2.35 | 0.94 | 0.40 – 2.25 |
| Very Low | 42.3 (134) | 1 |  | 1 |  |
| **Financial situation** |  |  |  |  |  |
| Worse off | 52.5 (32) | 1.97 * | 1.13 – 3.44 | 1.73 | 0.89 – 3.34 |
| Not worse off | 35.9 (102) | 1 |  | 1 |  |
| **Care setting** |  |  |  |  |  |
| Secondary | 34.2 (76) | 0.68 | 0.44 – 1.05 | 0.84 | 0.50 – 1.39 |
| Primary | 43.5 (60) | 1 |  | 1 |  |

| Diagnosis |  |  |  |  |  |
| --- | --- | --- | --- | --- | --- |
| Not recorded | 27.1 (13) | 0.85 | 0.42 – 1.72 | 0.80 | 0.35 – 1.80 |
| Other SMI | 30.4 (7) | 0.998 | 0.39 – 2.56 | 1.01 | 0.37 – 2.80 |
| Bipolar disorder | 56.7 (59) | 2.99** | 1.82 – 4.92 | 3.88** | 2.13 – 7.08 |
| Psychosis | 30.5 (57) | 1 |  | 1 |  |

a. Percentages are row percentages. \* $p < .05$ , \*\* $p < .001$ . Outcome variables is 'using the Internet a lot' vs 'Just a little or not at all'.

Participants who self-reported a decline in their mental health since the beginning of the pandemic were almost twice as likely to have used the Internet ‘a lot’ during the pandemic, compared to those that did not self-report a decline (Table 4).

**Table 4.** Association of health variables with Internet use.

|  | Using the Internet “a lot”<br>- % (N)<sup>a</sup> | Univariate Model |  | Multivariate Model |  |
| --- | --- | --- | --- | --- | --- |
|  |  | OR | 95% CI | Adj. OR | 95% CI |
| **Physical health** |  |  |  |  |  |
| Decline | 45.3 (53) | 1.55 | 0.99 – 2.44 | 1.39 | 0.84 – 2.30 |
| No decline | 34.8 (81) | 1 |  | 1 |  |
| **Mental health** |  |  |  |  |  |
| Decline | 46.9 (69) | 1.97* | 1.27 – 3.04 | 1.92* | 1.15 – 3.19 |
| No decline | 31.1 (64) | 1 |  | 1 |  |
| **Wellbeing (Mean, SD)** | 22.23 (8.46) | 0.98 | 0.96 – 1.01 | 1.01 | 0.98 – 1.04 |

a. Percentages are row percentages. \* $p < .05$ , \*\* $p < .001$ . Outcome variables is 'using the Internet a lot' vs 'Just a little or not at all'.

### Post-hoc analysis

To further explore the association between decline in mental health and greater use of the Internet during the pandemic restrictions, we examined whether decline in mental health was associated with any of the sample characteristics that were associated with Internet use. There was a significant association with diagnosis (x^2^(3) = 8.70, *P* = .03). More people with bipolar disorder self-reported a mental health decline (51.9%) compared to people with psychosis (34.8%) There was no association between decline in mental health and age (x^2^(3) = 2.99, *P* = .39).

In the light of this, we examined whether a decline in mental health was associated with greater use of the Internet after adjusting for age and diagnosis. We used a binary logistic regression, with age, diagnosis, and changes in mental health as independent variables, and use of the Internet as the outcome variable. Decline in mental health was still significantly associated with use of the Internet (*P* = .04, see Table 5), suggesting that decline in mental health still explained a unique portion of variance in use of the Internet after considering people’s age and diagnosis.

**Table 5.** Post-hoc exploratory analysis: Association of mental health decline with Internet use, adjusting for age and diagnosis.

|  | Using the Internet “a lot” - % (N)<sup>a</sup> | Univariate Model |  | Multivariate Model |  |
| --- | --- | --- | --- | --- | --- |
|  |  | OR | 95% CI | Adj. OR | 95% CI |
| **Age** |  |  |  |  |  |
| 18-30 | 54.9 (28) | 3.59* | 1.70 – 7.60 | 5.75** | 2.49 – 13.28 |
| 31-45 | 46.4 (45) | 2.55* | 1.34 – 4.87 | 3.23* | 1.57 – 6.67 |
| 46-65 | 31.9 (43) | 1.38 | 0.74 – 2.57 | 1.72 | 0.87 – 3.40 |
| 66+ | 25.3 (20) | 1 |  | 1 |  |
| **Diagnosis** |  |  |  |  |  |
| Not recorded | 27.1 (13) | 0.85 | 0.42 – 1.72 | 0.81 | 0.38 – 1.74 |
| Other SMI | 30.4% (7) | 0.998 | 0.39 – 2.56 | 0.87 | 0.31 – 2.47 |
| Bipolar disorder | 56.7 (59) | 2.99** | 1.82 – 4.92 | 3.92** | 2.24 – 6.87 |
| Psychosis | 30.5 (57) | 1 |  | 1 |  |
| **Mental health** |  |  |  |  |  |
| Decline | 46.9 (69) | 1.97* | 1.27 – 3.04 | 1.63* | 1.02– 2.62 |
| No decline | 31.1 (64) | 1 |  |  |  |

a. Percentages are row percentages. \* $p < .005$ , \*\* $p < .001$ . Outcome variables is 'using the Internet a lot' vs 'Just a little or not at all'.

## DISCUSSION

Most people with SMI were limited or non-users of the Internet during the pandemic restrictions. Although most participants were not affected by lack of Internet or device access, an important minority did not have access to the internet/ devices, and around half reported some skills deficiencies. Older adults with psychosis were the least likely to be regular Internet users during the pandemic.

Compared to findings among people with SMI prior to the pandemic, we found a much lower rate of non-users of the Internet (39.5% vs 85.6% in Tobitt, et al., 2019), but also a lower rate of frequent users of the Internet (37.1% vs 55% in Robotham, et al., 2016). Ownership of a digital device was high before the pandemic (Robotham, et al., 2016: 60% owned a computer; Hidalgo-Mazzei, et al., 2019: 67.8% owned tablet and 92.8% a smartphone) and remained as such in this study (79.8% owned a tablet or smartphone). It appears that the main change from pre-COVID to now is that more people are using the Internet. However, these differences should be interpreted with caution, considering sampling variations among the studies. For example, Tobitt, et al. (2019) recruited people with psychosis in community rehabilitation services (and therefore potentially more profoundly affected by their SMI), while Hidalgo-Mazzei, et al. (2019) recruited people with bipolar disorder that subscribed to an e-newsletter (thus potentially more confident with using online services). To minimise such bias in our study, participants had the option to complete the survey either online or offline (over the phone or with a hardcopy).

Worryingly, there seems to be a wide divide between those with SMI and the general population in terms of use of the Internet and self-reported internet skills (Lloyds Bank, 2021). During the pandemic restrictions, 5% of the UK population was off-line (compared to 39.5% in this study), and 85% of the UK population reported feeling confident in using the Internet (compared to 48.8% reporting outstanding or good knowledge about the Internet here).

The most common barriers for using the Internet reported here (e.g., lack of interest, difficulty of the Internet, and security/privacy concerns) have also been reported by SMI studies before the pandemic (lack of knowledge, skills, or understanding: Greer, Robotham, Simblett, Curtis, Griffiths and Wykes, 2019;Robotham, et al., 2016;Tobitt, et al., 2019) and in the general population during the pandemic (worry over privacy and security and finding the Internet too complicated or not interesting: Lloyds Bank, 2021). Although lack of interest might demonstrate an informed choice to not use the Internet, it might also mask deficits in skills and knowledge (French, Quinn and Yates, 2019). Despite 48% of our sample residing in areas of high or very high socioeconomic deprivation, financial barriers were reported by only 8% of our sample. Financial barriers have been more prominently reported in previous studies of SMI (Robotham, et al., 2016). This might be explained, up to an extent, by prices in mobile data falling steeply lately (Ofcom2020c).

Out of all the reported barriers, finding the Internet not useful was the most rarely reported (3.1%). This is positive, suggesting that most limited or non-users of the Internet recognised the benefits of engaging with the online world. This is further corroborated by the fact that almost 60% of the participants who reported a gap in their knowledge about the Internet expressed an interest in learning more about it.

Most performed activities online were accessing information and entertainment, staying in touch with friends and purchasing goods, probably due to restrictions in visiting other people or shops and spending more time in house. Least common activity was staying in touch with colleagues from work, probably since 80% of our sample was not in employment or furloughed during the pandemic.

Older people and those with a psychosis-spectrum disorders were more likely to be limited or non-users of the Internet during the pandemic. This is not surprising as older age is traditionally associated with less Internet engagement both in people with SMI (Robotham, et al., 2016;Tobitt, et al., 2019) and in the general population (National Health Service Digital [NHS Digital], 2019;Ofcom 2020a). However, during the pandemic, older adults were considered at-risk for experiencing severe effects of COVID-19 and were, therefore, advised to self-isolate and not leave their premises for long periods of time. For some of them, lack of Internet engagement might have meant lack of access to essential services and support. As such greater emphasis is needed in supporting older adults with SMI to use the Internet, as well ensuring offline access remains available. Regarding the role of diagnosis, barriers to using the Internet related to reduced concentration, hallucinations, or paranoid ideas; (Greer, et al., 2019;Schrank, Sibitz, Unger and Amering, 2010) might be more common in people with psychosis spectrum disorders than bipolar. In our sample, more people with bipolar disorder (27.6%) than psychosis (12.9%) were active in paid employment during the pandemic restrictions (working full or part-time and not being currently furloughed), so this might have been another reason for greater use of the Internet among people with bipolar.

This study demonstrated that participants who self-reported a decline in their mental health since the beginning of the pandemic restrictions, also reported using the Internet a lot, regardless of people’s age or diagnosis. This seems in agreement with reports in the general UK population that excessive consumption of COVID-19 related news in social media was associated with increased depression and anxiety (although the directionality of the association was not determined; Neill, Blair, Best, McGlinchey and Armour, 2021). However, we cannot be certain that the people who were using the internet a lot in our study were using it to access COVID-19 related news, although accessing information and entertainment was reported as the most common online activity.

Furthermore, it might be that people whose mental health declined used the Internet more intensively as a coping mechanism. For example, during the COVID-19 pandemic, about half of the Internet users in the UK reported using the Internet to support their mental and physical health and to feel less lonely (Lloyds Bank, 2021). This is a complex relationship that requires further investigation in terms of the online activities people were engaging with, their motivations and expectations from these activities, and the impact on their mental health. Qualitative work could be useful in further exploring these issues.

This study draws strength from its representative sample covering a wide range of diagnoses, geographic and socioeconomic areas, care settings, and ethnic backgrounds. However, the OWLS survey was kept to the shortest possible length to reduce participant burden. As a result, some of the reported variables were measured with single self-report items (e.g., have you used the internet a lot during the pandemic?) rather than more objective and fine-grained indicators (e.g., a complete break-down of activities with frequency and duration of engagement). For the same reasons we explored the common barriers found in the general population, but we did not ask our participants about barriers more specific to SMI. Despite this, this study provides important insights in use of the Internet by people with SMI during the pandemic restrictions, however, further exploration is warranted to understand more clearly some of the associations we identified.

Overall, although people with SMI may have become more digitally engaged since the pandemic began, there is still a wide gap in Internet use between people with SMI and people without SMI. However, this appears to be mainly driven by lack of skills or interest, rather than lack of Internet or device access. Digital skills among people with SMI should be further explored to understand the main areas of deficit. Digital inclusion efforts for people with SMI should focus not only on providing people with devices but also offering training and support to improve skills. It is of concern that a vulnerable sub-group (older people with psychosis) appear to be at greater risk for digital exclusion during the pandemic. Digital inclusion interventions should focus on barriers related to older age and a diagnosis of psychosis.

## Supporting information

STROBE checklist

## Data Availability

Data can be made available from authors on request.

https://osf.io/4qwr7/?view_only=32224b0193a54a09a6f9eee0873351a7

## ACKNOWLEDGEMENTS

We thank the participants in the OWLS study and NHS mental health staff for their support with this study.

